# An Epidemiological Study of the Impact of Doula Care on Black Birth Outcomes in Baltimore, Maryland

**DOI:** 10.64898/2026.09.30.26364355

**Authors:** Elizabeth A. Watson

## Abstract

This study examines the impact of various maternal factors and continuous labor support on birth outcomes among 124 birthing people in the Baltimore region of Maryland who sought and/or received doula care. The cohort primarily consists of Black birthing people and includes detailed data on maternal age, race, education level, employment status, health insurance status, income level, preexisting health challenges, and pregnancy or delivery-related health challenges. The study also explores whether the pregnancy was the birthing person’s first, as well as neonatal outcomes such as NICU admission, low birth weight, and other newborn health concerns. Due dates and delivery dates were compared and analyzed, where available, to provide a comprehensive understanding of the timing of births among the cohort. The findings highlight known correlations between maternal socioeconomic factors, health factors, and adverse birth outcomes, emphasizing the critical role of doula support during the prenatal period and labor and delivery in improving maternal and neonatal health, as well as in providing culturally competent care and positive birth experiences. This research contributes to the growing body of evidence on the benefits of doula care, particularly for marginalized communities, and underscores the need for targeted interventions to address health disparities among Black Birthing persons.

## I. Introduction

Maternal and neonatal health and birth outcomes are significant indicators of the health of a population, and yet many disparities exist, particularly among Black and other minority birthing people. It is a public health crisis that birth outcomes contributing to infant mortality present disparities that are greater than they were during antebellum slavery, despite advances in obstetric care, maternal healthcare, and neonatal care technology [1]. In the state of Maryland, most recent data shows Black Marylanders had a lower percentage of the state’s total births – 30% compared to 41% for white Birthing persons – and yet significantly higher rates of teen births (22.9 per 1,000), preterm births (13% of all births), low birth weight (13% of all births), and c-section deliveries (39% of all births) [2]-[3].

Multiple studies over the last decade have examined potential causes of these disparities, as well as practical solutions. Giscombe and Lobel [4] examined the effects of stress on pregnancy experiences, maternal health, infant health and birth outcomes. Their research suggests that Black birthing people encounter greater base levels of stress as well as chronic stress in the form of racism throughout their lives, and the cumulative effect of this stress has significant negative impacts on their health and the health of their pregnancies.

While the study examined sources of stress including socioeconomic and health-related factors, such stressors are ultimately part of the vicious circle of racism, highlighting the systemic source of the large disparities affecting Black Birthing persons and infants. A more recent study found that between a fifth and a third of minority respondents experienced racial discrimination in a healthcare setting, and up to 22% avoid care due to concerns of experiencing racial discrimination [1]. In a country where Black birthing people are more likely than white birthing people to receive late prenatal care – or no prenatal care at all – and have significantly higher rates of cesarian deliveries in low-risk pregnancies, this suggests that one of the main barriers to care for Black birthing people is race-based discrimination, adding to the cumulative stress of lived experiences of racism [5].

Socioeconomic factors have also been shown to be associated with birth outcomes but have proven difficult to measure and interpret in accurate and meaningful ways. The socioeconomic factors studied often contribute to a poorly defined “socioeconomic class,” and often only one side of the equation is examined, such as either paternal or maternal education and income levels, but not both [6]. Unfortunately, continuous labor support – such as doula care – is not reimbursed by Medicaid or other insurance providers and is often inaccessible to minority or socioeconomically disadvantaged communities and populations. A study by Kozhimannil et al. [7] showed that introducing Medicaid reimbursement for doula services, even at a rate of $300 per birth, could be cost effective, significantly reducing the cost of c-section deliveries and the cost burden of caring for premature infants needing intensive care or maternal health conditions. As this study will highlight, this is due to the advocacy and care that doulas provide, which reduces the rate of unnecessary c-sections through advocacy and education as well as reducing the medically necessary c-section rate and premature births through their prenatal care.

Overall, factors like family poverty, parental education, and parental employment status have been shown to be associated with low birth weight and preterm birth. Unfortunately, low birth weight has been shown to increase the risk of adulthood diseases such as cardiovascular disease, gestational diabetes, hypertension, and type 2 diabetes—all of which occur at higher rates among the Black community already [8]-[10].

This increased risk association remains clear even after studies have adjusted for lifestyle factors and other social determinants of health [11], suggesting that the vicious circle of racism is perpetuating birth outcome and infant health disparities among Black Birthing persons.

In response to these disparities and concerns over culturally competent pregnancy care that preserves the autonomy of birthing persons, increasing cesarean delivery rates, and rising healthcare costs associated with pregnancy, labor and delivery, interest in doula care has increased. Research has shown that implementing Medicaid reimbursement rates for doula care – which would make continuous labor support through doulas more accessible and affordable – could result in savings to Medicaid programs and ultimately reduce the percentage of GDP spent on healthcare in the US related to c-section rates, low birth weight, and other complications or negative birth outcomes [7].

This study of birthing persons in the Baltimore region of Maryland, all of whom sought doula care, is best interpreted within the context of existing research on maternal health, birth outcomes, and neonatal health as indicators and potential risk factors for future health. It is hypothesized that doula care is associated with reduced c-section delivery rates as well as reduced preterm birth rates.

## II. Methodology

This study uses an open cohort design and descriptive statistics to examine maternal health and socioeconomic factors and birth outcomes among 124 people in the Baltimore region of Maryland, who completed, at minimum, an intake process with MOMCares, MD, between 2022 and 2024.

MOMCares has kept intake data, appointment data, and post-birth surveys for patients between 2022 and early June 2024. The cohort consists of a total of 124 birthing people, some of whom ultimately received doula care and services through MOMCares and some who did not. Some data points are missing for some patients. A maternal and neonatal demographics and health survey was sent via email to each member of the cohort, to confirm existing data that was not clear and to fill in missing data points wherever possible. This survey asked straightforward questions about each of these data points, as well as if doula care contributed to a more culturally competent and positive birth experience. A copy of the survey is available in the appendix; the survey response rate was 12%.

Data was collected through a combination of this structured survey and electronic health records kept by MOMCares. The data collected focused on: maternal age, race, employment status, income level, education, health insurance status, maternal health conditions (whether preexisting, pregnancy related, or delivery related), if it was the birthing person’s first baby, delivery method, low birth weight or not, NICU admission, newborn health concerns, and due dates and actual delivery dates to ascertain preterm births.

Data was compiled into one excel table for analysis, uploaded into SAS OnDemand (SAS 3.81 Enterprise Edition), and converted into a SAS format for analysis. Descriptive statistics were used to summarize and compare the data. The analysis included frequencies and percentages, measures of central tendency and dispersion, and cross-tabulations.

Pregnancy Risk Assessment Monitoring System (PRAMS) data for the Maryland site from phase 8 (2016-2021) was obtained and used for comparison to the MOMCares cohort. PRAMS is a survey of maternal and infant health sent to birthing people who recently gave birth to a living neonate. It has been conducted since 1987 by the Centers for Disease Control and Prevention, and each site samples anywhere between 1,000 and 3,000 birthing people per year, with each jurisdiction of the site sampling between 100 and 250 birthing people. Surveys are conducted by mail or telephone interview each month, to birthing people within 2 to 4 months after delivery of a live birth. Some groups of birthing people are sampled at higher rates, to ensure adequate data collection that is representative of the population sampled [12]. Once access to deidentified PRAMS phase 8 data was granted, the data file was filtered to include only the records with the ID prefix “2016MD.” Moving forward in discussion of the methodology, data analysis, results, and discussion, this is the PRAMS data that was utilized for comparison to the MOMCares cohort.

Simple random sampling was used on the Maryland-site PRAMS data to draw a sample that is racially similar to the MOMCares sample for whom race, and delivery method data is available. This smaller PRAMS sample and the MOMCares sample each consisted of 18 Black birthing people who gave birth in various ways. We ran Fisher’s exact tests (significance threshold of *p* < 0.05) to determine the similarity between the two samples among race (*p* = 0.2429) and delivery method (*p* = 0.0973). The results indicate that the two samples are comparable in these selected categories.

Although the cohort consisted of 124 birthing people in total (*N*), Table 1 shows, for how many people of the cohort (*n*) there is data for each variable. The variables for which there was the least data were delivery date, the Birthing Person’s age at delivery, if the baby had low birth weight, the delivery method, and the length of hospital stay. The categories of prenatal and postpartum appointments attended have low amounts of participation because while all 124 birthing people applied for services through MOMCares, and therefore had an intake form, not all of them were eligible to receive services. The 32 birthing people who attended prenatal appointments in some measure are the only members of the total cohort who received doula care through MOMCares between 2022 and June of 2024.

**TABLE I.**
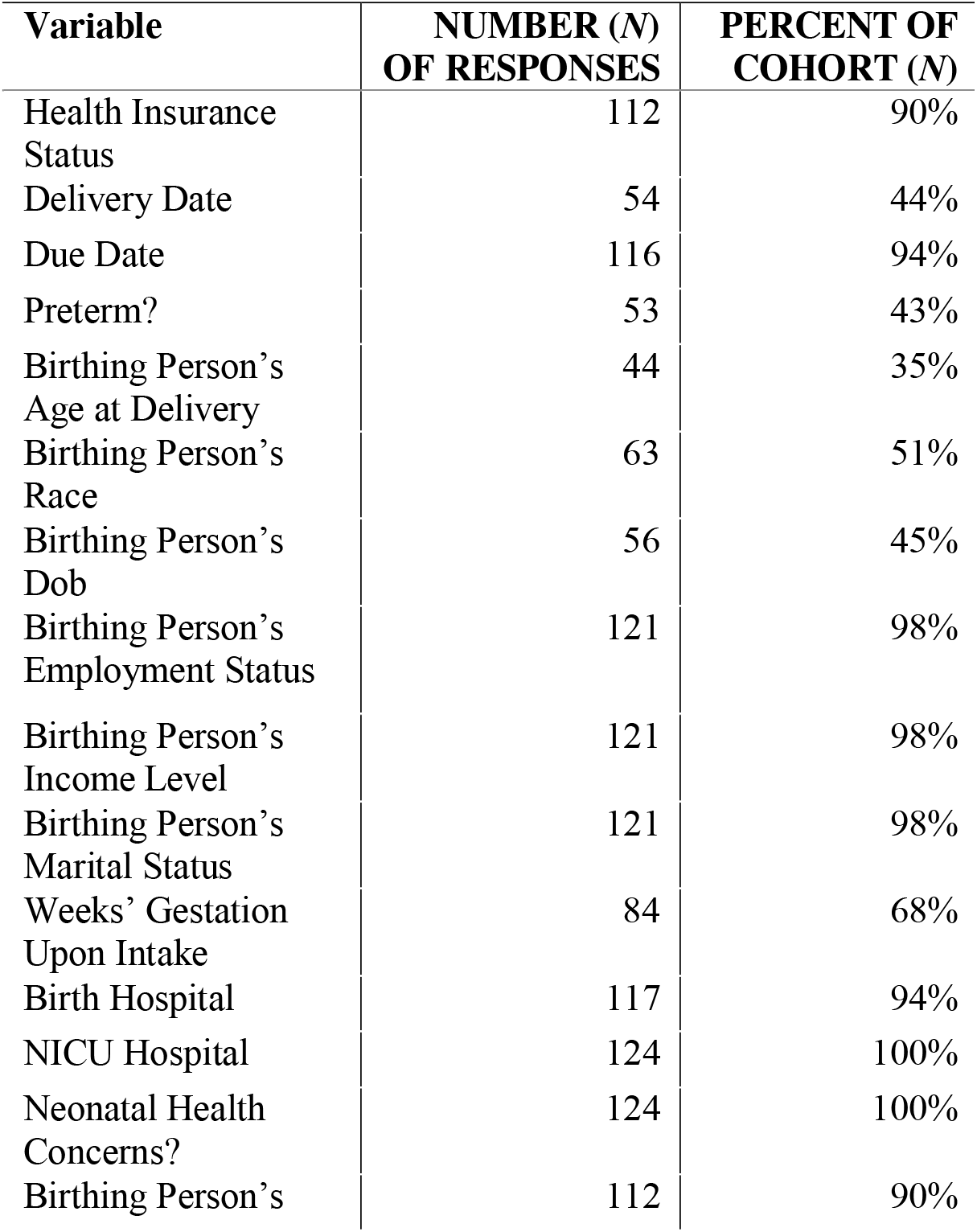
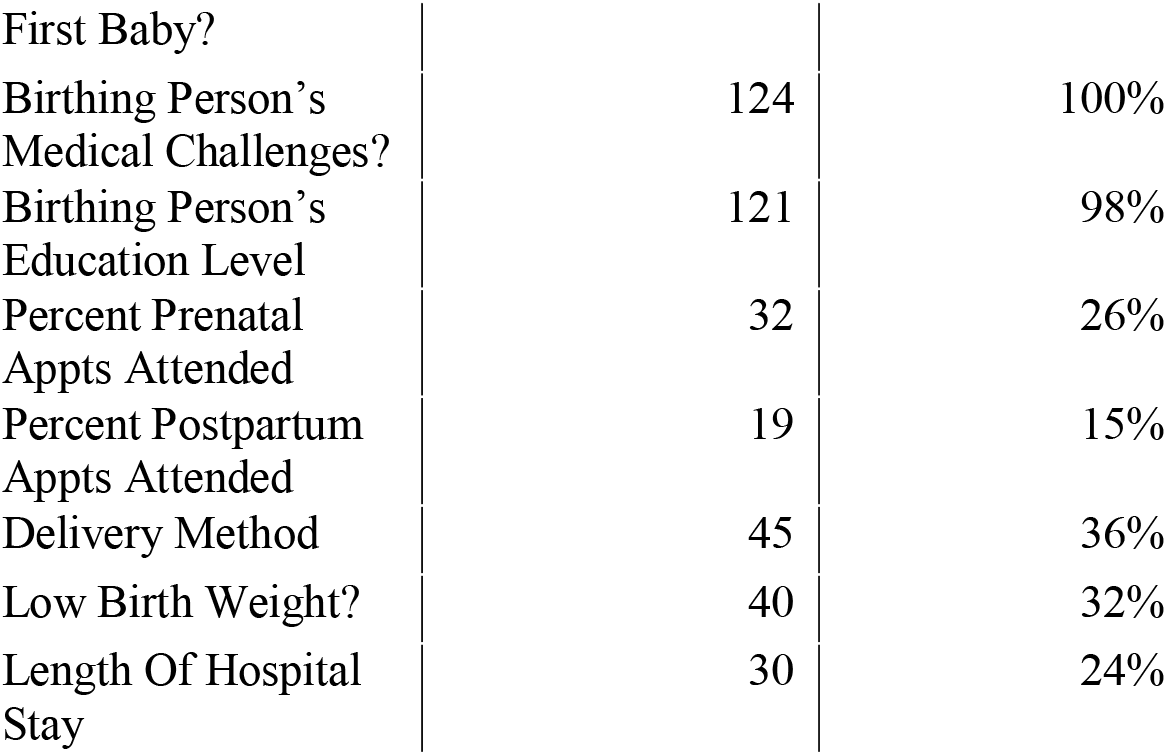
Showing the variables examined and for what percentage of the cohort responses were obtained.

The MOMCares sample (*N* = 124) primarily consisted of Black birthing people (race *n* = 63), with that race making up 84.13% of the cohort for which racial data exists. 88.39% of the cohort (health insurance status *n* = 112) were enrolled in Medicaid, while 11.61% were enrolled in private insurance through their employer. 34.71% (birthing person’s employment status *n* = 121) were employed full time, while 23.41% were unemployed and intending to stay home. As to income level, 54.55% (birthing person’s income level *n* = 121) earned less than $20,000 annually; 24.79% earned between $20,000 to $34,999. 72.75% of the cohort (birthing person’s marital status *n* = 121) were single at the time of delivery. 28.1% (birthing person’s education level *n* = 121) had graduated high school, and 35.54% had taken some college courses but not completed a degree. The average age of birthing people in this cohort (birthing person’s age *n* = 42) was approximately 29. 29.91% of the cohort (birth hospital *n* = 117) chose Mercy Medical Center as their birthing hospital. Tables 2 through 4 show the complete SAS output for these descriptive statistics.

**TABLE II.** Mean and range (measure of central tendency) for variable “birthing person’s age at delivery.”

| Birthing person's age at delivery |  |  |  |  |
| --- | --- | --- | --- | --- |
| N | Mean | Std Dev | Minimum | Maximum |
| 44 | 28.9090909 | 5.5649158 | 17.0000000 | 38.0000000 |

**TABLE III.** Showing the distribution of births in the cohort among hospitals or hospital systems.

| Birth Hospital | Frequency | Percent |
| --- | --- | --- |
| Anne Arundel Medical Center | 2 | 1.71 |
| Birthing Center | 1 | 0.85 |
| Franklin Square Hospital | 6 | 5.13 |
| Greater Baltimore Medical Center | 9 | 7.69 |
| Harbor Hospital | 3 | 2.56 |
| Holy Cross German Town | 2 | 1.71 |
| Home Birth | 3 | 2.56 |
| Howard County General | 1 | 0.85 |
| Johns Hopkins Medical Center | 15 | 12.82 |
| Mercy Medical Center | 35 | 29.91 |
| N/A | 1 | 0.85 |
| Shady Grove | 1 | 0.85 |
| Sinai Hospital | 6 | 5.13 |
| St. Agnes | 4 | 3.42 |
| St. Joseph Medical Center | 13 | 11.11 |
| University Of Florida Health Jacksonville | 1 | 0.85 |
| University Of Maryland Medical Center | 11 | 9.40 |
| White Oak Medical Center | 1 | 0.85 |
| Women's Birth Center Holistic | 1 | 0.85 |
| Foreign Born | 1 | 0.85 |

**TABLE IV.**
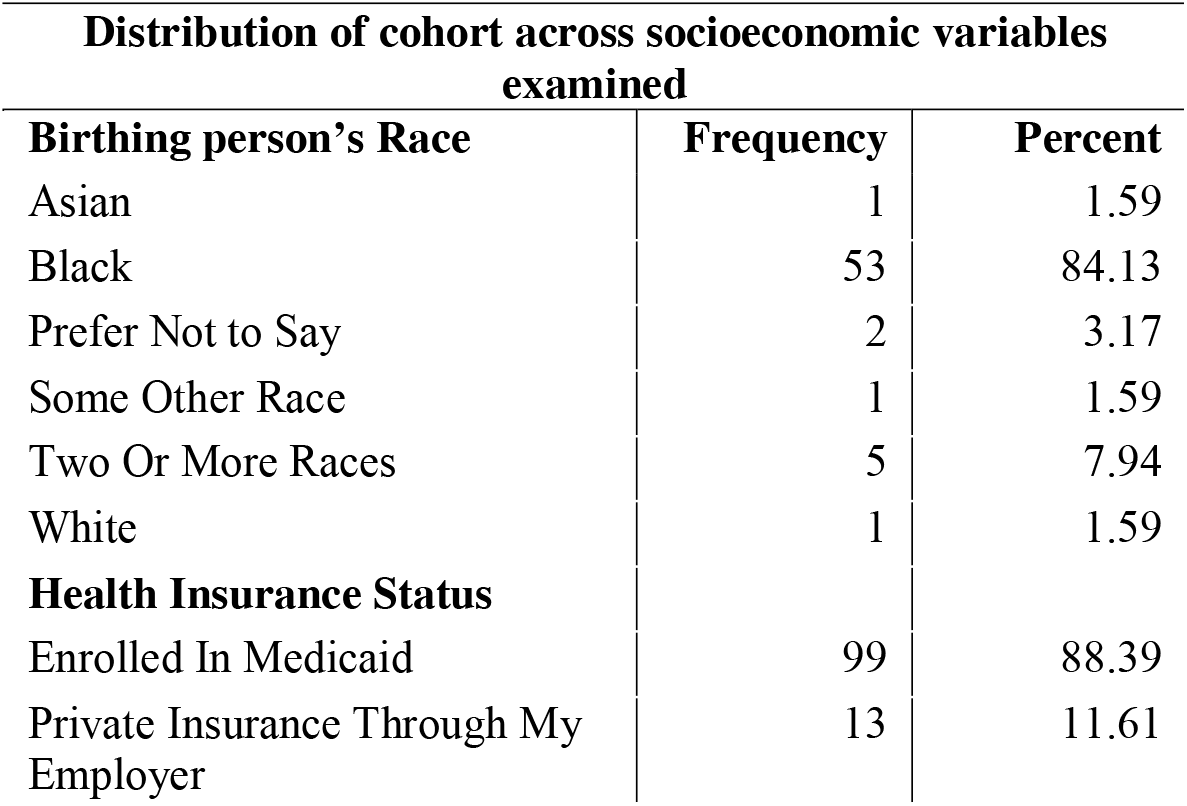
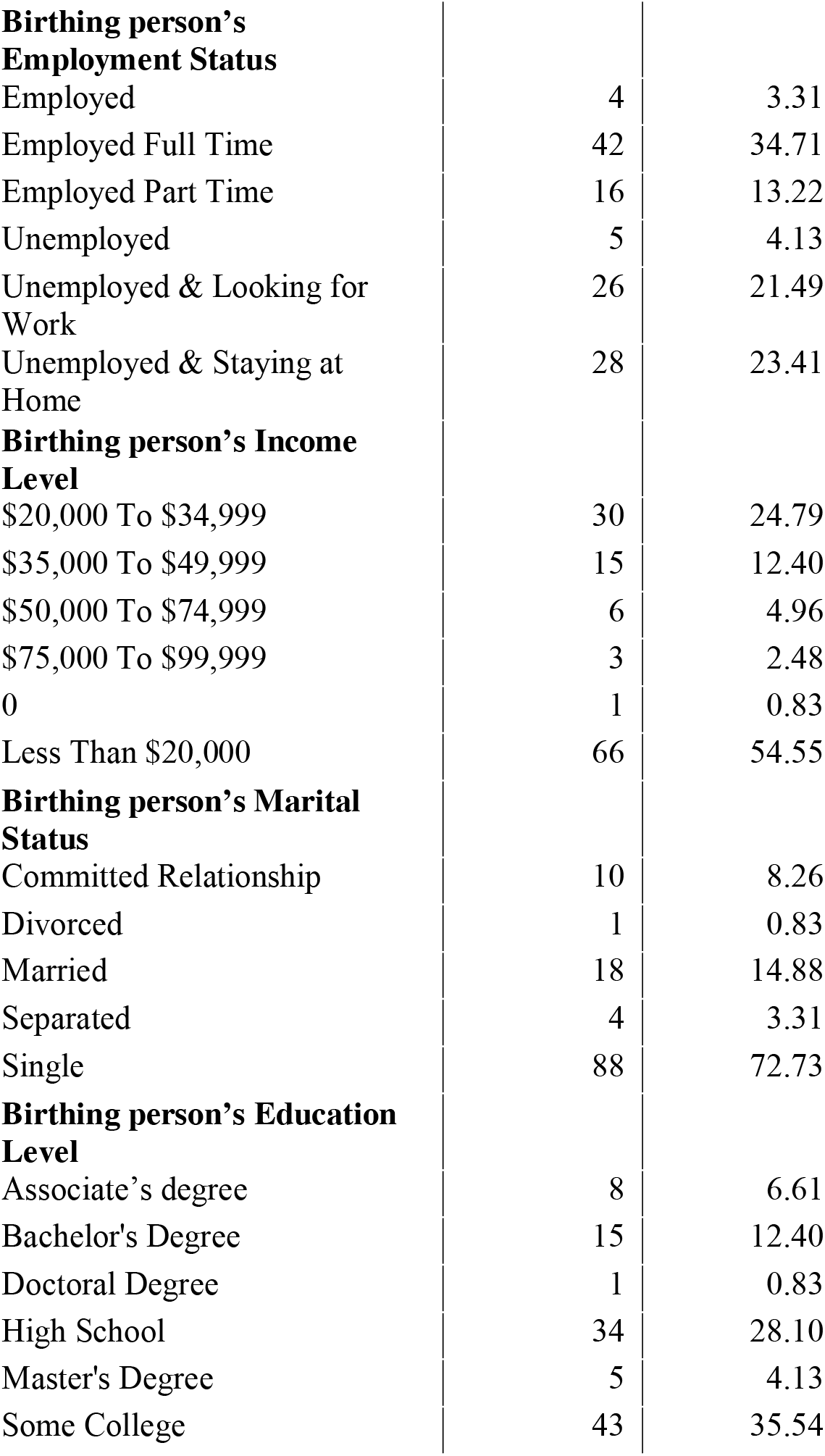
Distribution of cohort across socioeconomic variables.

| Distribution of cohort across socioeconomic variables examined |  |  |
| --- | --- | --- |
| Birthing person's Race | Frequency | Percent |
| Asian | 1 | 1.59 |
| Black | 53 | 84.13 |
| Prefer Not to Say | 2 | 3.17 |
| Some Other Race | 1 | 1.59 |
| Two Or More Races | 5 | 7.94 |
| White | 1 | 1.59 |
| **Health Insurance Status** |  |  |
| Enrolled In Medicaid | 99 | 88.39 |
| Private Insurance Through My Employer | 13 | 11.61 |
| **Birthing person's Employment Status** |  |  |
| Employed | 4 | 3.31 |
| Employed Full Time | 42 | 34.71 |
| Employed Part Time | 16 | 13.22 |
| Unemployed | 5 | 4.13 |
| Unemployed & Looking for Work | 26 | 21.49 |
| Unemployed & Staying at Home | 28 | 23.41 |
| **Birthing person's Income Level** |  |  |
| \$20,000 To \$34,999 | 30 | 24.79 |
| \$35,000 To \$49,999 | 15 | 12.40 |
| \$50,000 To \$74,999 | 6 | 4.96 |
| \$75,000 To \$99,999 | 3 | 2.48 |
| 0 | 1 | 0.83 |
| Less Than \$20,000 | 66 | 54.55 |
| **Birthing person's Marital Status** |  |  |
| Committed Relationship | 10 | 8.26 |
| Divorced | 1 | 0.83 |
| Married | 18 | 14.88 |
| Separated | 4 | 3.31 |
| Single | 88 | 72.73 |
| **Birthing person's Education Level** |  |  |
| Associate's degree | 8 | 6.61 |
| Bachelor's Degree | 15 | 12.40 |
| Doctoral Degree | 1 | 0.83 |
| High School | 34 | 28.10 |
| Master's Degree | 5 | 4.13 |
| Some College | 43 | 35.54 |

## III. Results

The aim of this data analysis is to determine if c-section, low birth weight, and preterm delivery weights are lower among birthing people who receive doula care. When using *p* values, the null hypothesis is assumed to be that there is no relationship between the compared variables (ex. H_0_ = there is no statistical association between maternal race and delivery method). A *p* value of 0.05 or lower will reject the null hypothesis. An inability to reject the null hypothesis does not indicate that a relationship between two variables does not exist, only that this study and sample cannot rule out that a significant relationship may not exist.

Among the 124 birthing-person MOMCares cohort, 24.44% of deliveries were via c-section, while 2.23% were successful vaginal delivery after c-section (VBAC), and 73.33% were vaginal deliveries (Table 5). Among the Black birthing people of the cohort, c-sections made up 29.41% of deliveries, while vaginal deliveries made up 70.59% (Table 6). Likely due to the small sample size for which data exists for both variables, a Fisher’s exact test on the comparison provided a *p* value of 0.3186. Thus, we are unable to reject the null hypothesis that race and delivery method are not related.

**TABLE V.** Distribution of delivery methods among the cohort.

| Delivery Method | Frequency | Percent |
| --- | --- | --- |
| C-Section | 11 | 24.44 |
| VBAC | 1 | 2.23 |
| Vaginal | 33 | 73.33 |

**TABLE VI.** Distribution of delivery methods among black birthing people in the cohort.

| Delivery Method | Births | Percent |
| --- | --- | --- |
| C-Section | 5 | 29.41 |
| Vaginal | 12 | 70.59 |
| Total | 17 | 100 |

Preterm births made up 15.09% of the births for which there is data among the cohort, while full-term births made up the other 84.91% of births (Table 7). Among the Black members of the cohort, preterm deliveries made up 22.73% of births, and full-term births made up 77.27% (Table 8). Again, a Fisher’s exact test returns a *p* value of 0.3830, too high to reject the null hypothesis that maternal race and preterm birth are not associated.

**TABLE VII.** Distribution of preterm vs. Full-term birth across cohort.

| Preterm | Frequency | Percent |
| --- | --- | --- |
| No | 45 | 84.91 |
| Yes | 8 | 15.09 |

**TABLE VIII.** Distribution of preterm births across black birthing people.

| Preterm | Frequency | Percent |
| --- | --- | --- |
| No | 17 | 77.27 |
| Yes | 5 | 22.73 |
| Total | 22 | 100 |

Low birth weight infants (infants weighing less than 2,500 grams at birth) were associated with 7.50% of births among the MOMCares cohort. Infants with a normal birth weight made up 92.50% of births (Table 9). Among the Black birthing people in the cohort, low birth weight infants were 14.29% of deliveries (Table 10). This comparison produced a *p* value of 0.6044 in a Fisher’s exact test, again indicating that we cannot reject the null hypothesis that low birth weight and maternal race are not associated.

**TABLE IX.** Distribution of low-birth-weight deliveries across cohort.

| Low Birth Weight | Frequency | Percent |
| --- | --- | --- |
| NO | 37 | 92.50 |
| YES | 3 | 7.50 |

**TABLE X.** Distribution of low-birth-weight deliveries among black birthing people in the cohort.

| Low Birth Weight (<2500g) | Frequency | Percent |
| --- | --- | --- |
| No | 12 | 85.71 |
| Yes | 2 | 14.29 |
| Total | 11 | 100 |

Pregnancy Risk Assessment Monitoring System (PRAMS) phase 8 (2016-2021) data for the Maryland site yielded comparison results. The overall c-section rate for this group was 36.51%, while for the Black members of the PRAMS cohort it was 40.24% (Tables 11 and 12). Once simple random sampling was used to create an 18 member cohort of Black PRAMS birthing people, similar analysis revealed that the c-section rate among this smaller cohort was 22.22% (Table 13). Fisher’s exact test between this simple random sample and the 18 members of the MOMCares cohort, for whom we had both maternal race and delivery method data, produced *p* values of 0.2140 for delivery method and 0.2429 for maternal race.

**TABLE XI.** PRAMS phase 8, Maryland delivery method distribution.

| Delivery Method | Frequency | Percent |
| --- | --- | --- |
| C-Section | 349 | 36.51 |
| VBAC | 24 | 2.51 |
| Vaginal | 583 | 60.98 |

**TABLE XII.** PRAMS phase 8, Maryland delivery method distribution among black birthing people.

| Delivery method | Frequency | Percent |
| --- | --- | --- |
| C-section | 136 | 40.24 |
| VBAC | 13 | 3.85 |
| Vaginal | 189 | 55.92 |

**TABLE XIII.**
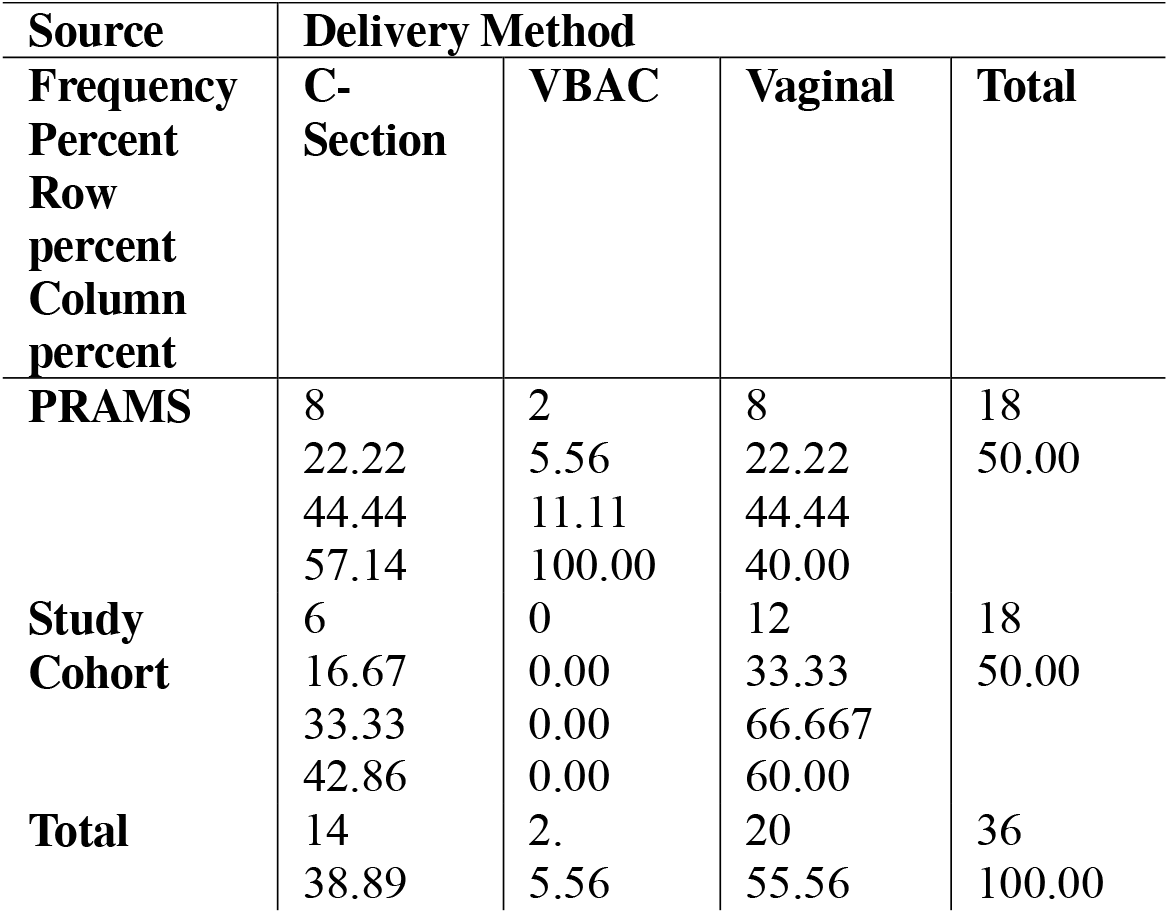
Delivery method cross-tabulation between cohort and PRAMS phase 8, Maryland.

These *p* values indicate that the two samples are similar enough to be comparable, as there is no statistically significant difference between them.

With regards to maternal health, conditions reported were categorized into delivery related conditions, pregnancy related conditions, pregnancy and delivery related conditions (ex. Group B strep, which is screened for and diagnosed during pregnancy but can be transmitted to the baby during delivery), preexisting conditions, and preexisting conditions which contributed to high-risk pregnancy and delivery (chronic hypertension, type 2 diabetes, and heart conditions, for example). 64.52% of the MOMCares cohort reported no maternal medical challenges whatsoever. 4.03% reported a delivery related condition, 5.65% reported a pregnancy related condition, 2.42% reported a condition which was associated with both pregnancy and delivery, 10.48% reported a preexisting condition unrelated to pregnancy or delivery, and 12.90% reported a preexisting condition that contributes to high risk pregnancy and delivery (Table 14).

**TABLE XIV.** Distribution of medical challenges across cohort.

| Medical Complication Category | Frequency | Percent |
| --- | --- | --- |
| Delivery Related Condition | 5 | 4.03 |
| N/A | 80 | 64.52 |
| Pre-Existing Condition | 13 | 10.48 |
| Pre-Existing Condition Contributing to High-Risk Pregnancy and Delivery | 16 | 12.90 |
| Pregnancy And Delivery Related Condition | 3 | 2.42 |
| Pregnancy Related Condition | 7 | 5.65 |

When comparing maternal health conditions (or lack thereof) to delivery methods, we discovered that among this cohort 25% of those with a delivery related condition had a c-section delivery, as well as 100% of those with a pregnancy related condition (Table 15). A Fisher’s exact text for this comparison yielded a *p* value of 0.0061, allowinf us to reject the null hypothesis that maternal health conditions and delivery method are not associated amongst the study cohort. 25% of those with a preexisting condition contributing to high risk pregnancy and delivery experienced preterm births. The other preterm births reported by the cohort primarily occurred in birthing people who reported no medical challenges (Table 16). The *p* value in a Fisher’s exact test for this comparison was 0.0615, again leaving us unable to reject the null hypothesis that maternal health and preterm births are not associated. However, this *p* value, less than 0.10, is only of marginal significance, and it’s worth examining such results in the broader context of previous research and known disparities.

**TABLE XV.**
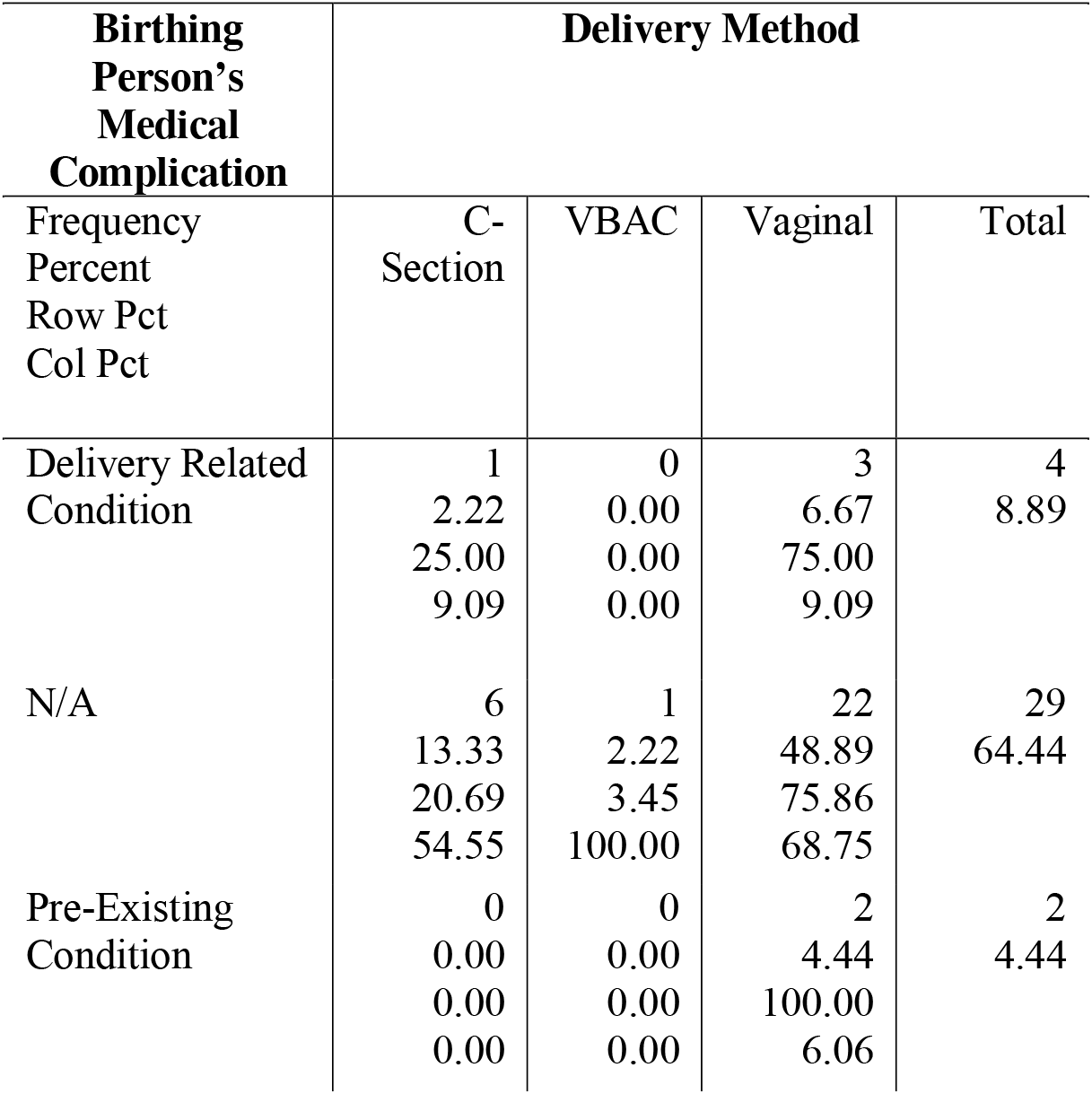
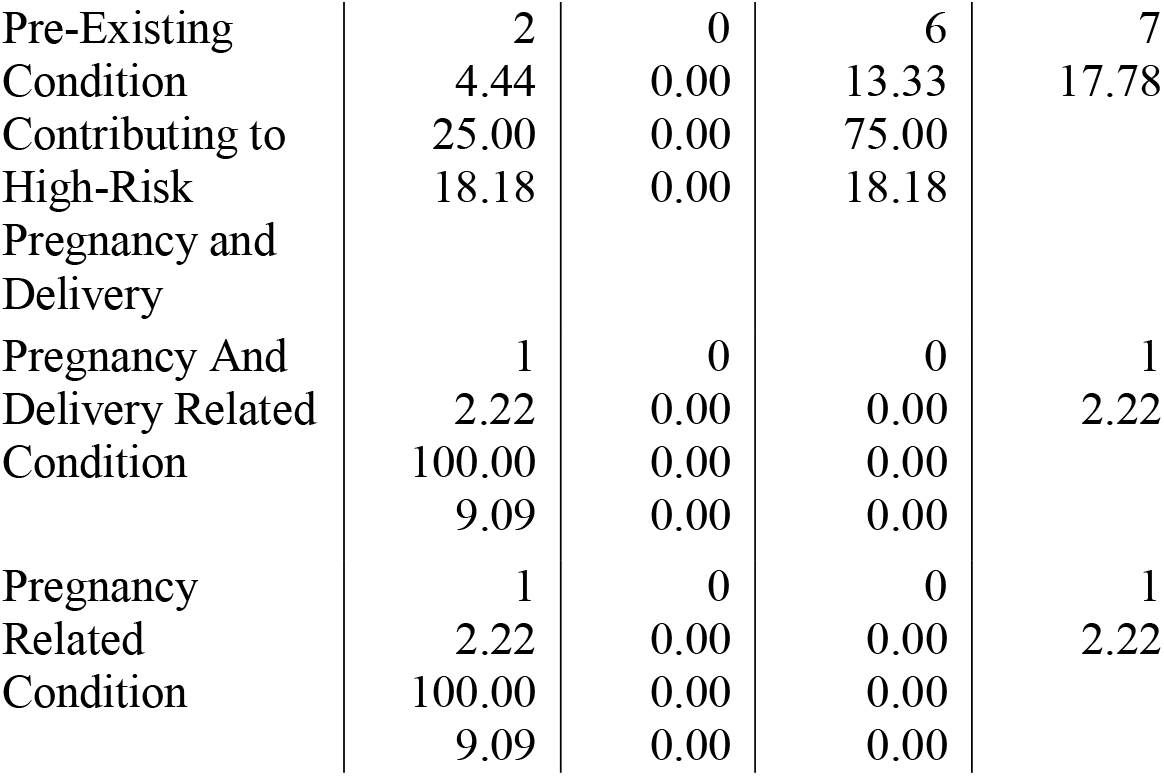
Distribution of delivery methods across medical complication categories among cohort.

**TABLE XVI.** Distribution of preterm births by birthing person’s medical complication category.

| Birthing person's medical complication category | Preterm |  |  |
| --- | --- | --- | --- |
|  | No | Yes | Total |
| Frequency<br>percent<br>row pct<br>col pct |  |  |  |
| Delivery related condition | 2<br>3.77<br>100.0<br>0<br>4.44 | 0<br>0.00<br>0.00<br>0.00 | 2<br>3.7<br>7 |
| N/a | 29<br>54.72<br>85.29<br>64.44 | 5<br>9.43<br>14.71<br>62.50 | 34<br>64.<br>15 |
| Pre-existing condition | 6<br>11.32<br>85.71<br>13.33 | 1<br>1.89<br>14.29<br>12.50 | 7<br>13.<br>21 |
| Pre-existing condition contributing to high risk pregnancy and delivery | 6<br>11.32<br>75.00<br>13.33 | 2<br>3.77<br>25.00<br>25.00 | 8<br>15.<br>09 |
| Pregnancy related condition | 2<br>3.77<br>100.0<br>0<br>4.44 | 0<br>0.00<br>0.00<br>0.00 | 2<br>3.7<br>7 |
| Pregnancy and delivery related condition | 0<br>0.00<br>.<br>0.00 | 0<br>0.00<br>.<br>0.00 | 0<br>0.0<br>0 |

Among the members of the MOMCares cohort who received doula care services, the c-section rate was 20%; among doula care patients who attended all of their prenatal appointments as opposed to a portion of them, this rate was 18.52% (Table 17). Using logistic regression, we found the regression coefficient for this relationship is -9.3588, while the odds ratio was calculated at <0.001, suggesting that as prenatal doula appointment attendance increases, the odds of c-section delivery decrease. Preterm births among this section of the cohort accounted for 10.34% of births; those who attended all of their prenatal appointments experienced preterm birth at a rate of 7.69% (Table 18). This comparison had a regression coefficient of -5.3429 and a odds ratio of 0.005, again suggesting that prenatal appointment attendance and preterm birth are negatively related – as prenatal appointment attendance increases, the odds of a preterm delivery decrease. Low birth weight deliveries represented 3.33% of deliveries among those who received doula care, and none of these occurred in birthing people who attended all of their prenatal doula appointments (Table 19). The regression coefficient for comparison of these two variables was calculated at -13.1795, with an odds ratio of <0.001 – suggesting a strong negative relationship, wherein the odds of low birth weight delivery decreases as attendance at prenatal doula appointments increases.

**TABLE XVII.**
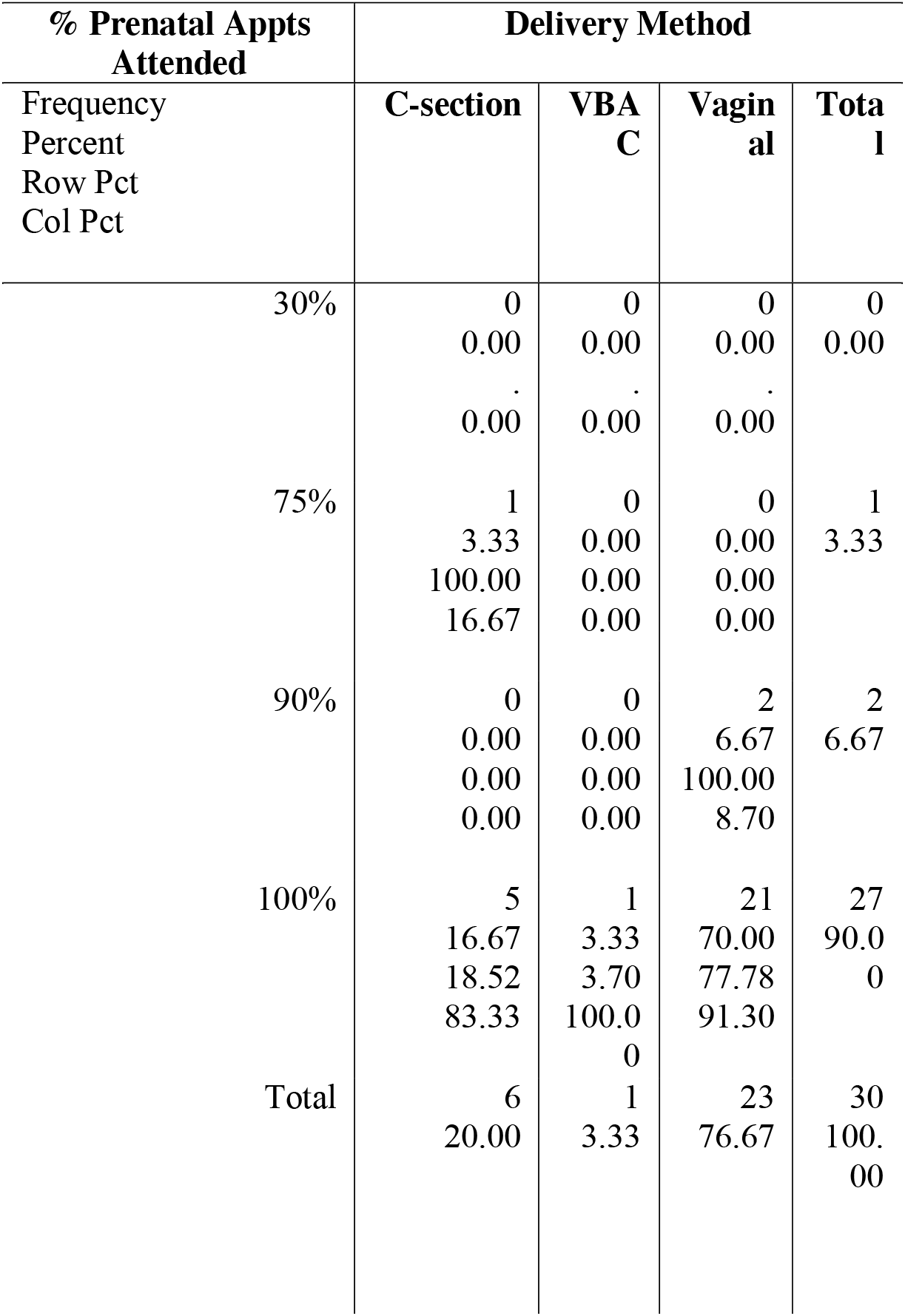
Distribution of delivery methods among doula care patients.

| **% Prenatal Appts Attended** | **Delivery Method** |  |  |  |
| --- | --- | --- | --- | --- |
| Frequency<br>Percent<br>Row Pct<br>Col Pct | **C-section** | **VBA C** | **Vaginal** | **Total** |
| 30% | 0<br>0.00<br>.00<br>0.00 | 0<br>0.00<br>.00<br>0.00 | 0<br>0.00<br>.00<br>0.00 | 0<br>0.00 |
| 75% | 1<br>3.33<br>100.00<br>16.67 | 0<br>0.00<br>0.00<br>0.00 | 0<br>0.00<br>0.00<br>0.00 | 1<br>3.33 |
| 90% | 0<br>0.00<br>0.00<br>0.00 | 0<br>0.00<br>0.00<br>0.00 | 2<br>6.67<br>100.00<br>8.70 | 2<br>6.67 |
| 100% | 5<br>16.67<br>18.52<br>83.33 | 1<br>3.33<br>3.70<br>100.00 | 21<br>70.00<br>77.78<br>91.30 | 27<br>90.0<br>0 |
| Total | 6<br>20.00 | 1<br>3.33 | 23<br>76.67 | 30<br>100.00 |

**TABLE XVIII.**
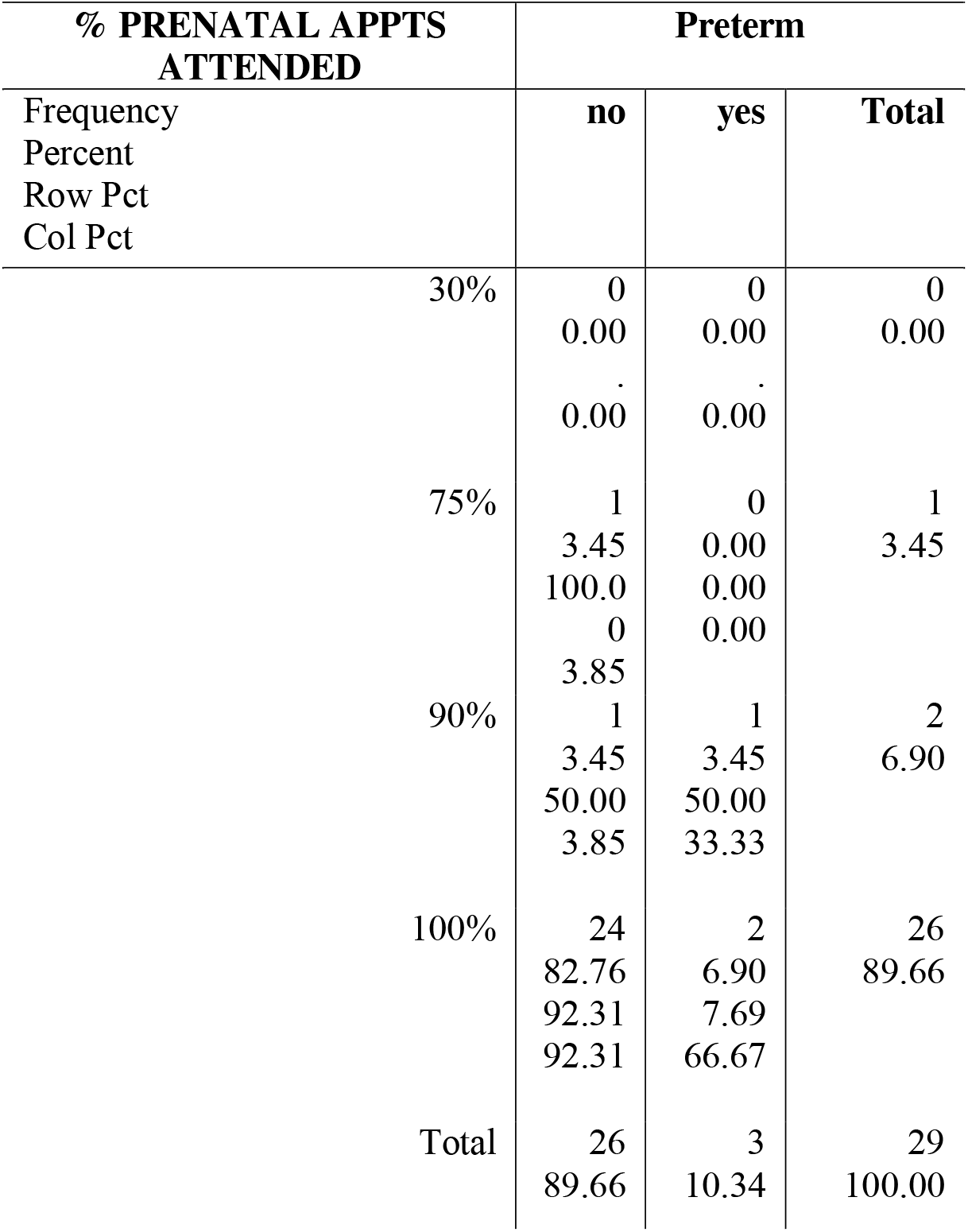
Distribution of preterm births across doula care patients.

**TABLE XIX.**
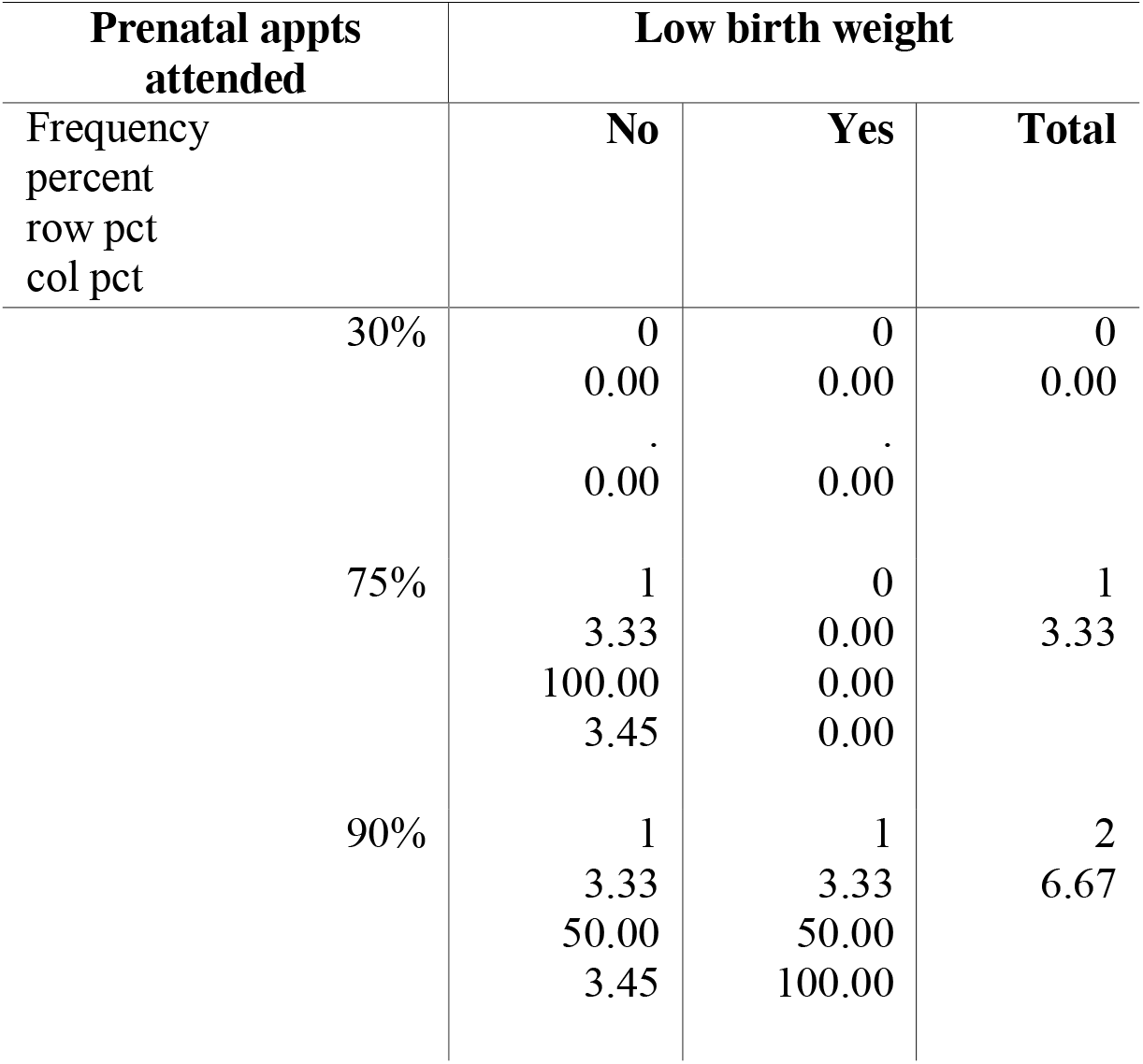
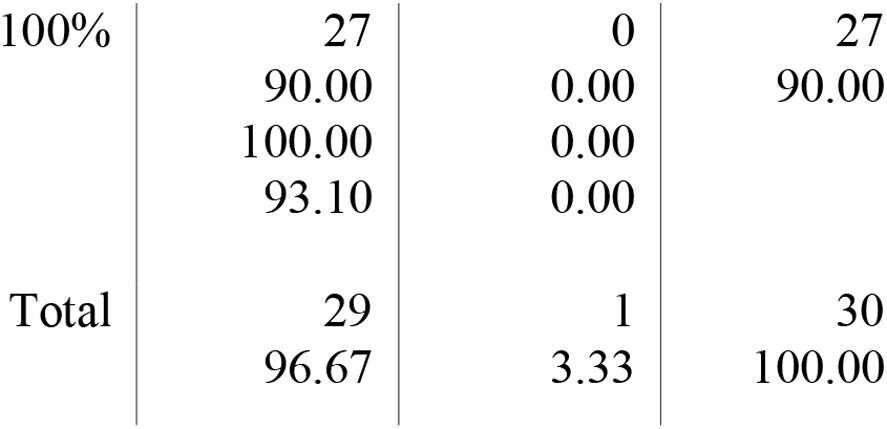
Distribution of low-birth-weight deliveries across doula care patients.

## IV. Discussion

Results of this population-based study suggest that continuous labor and prenatal support through a doula lowers the risk of c-section delivery, preterm delivery, and low birth weight. Significantly lower rates of c-section and preterm delivery, as well as low birth weight, were observed among those members of the cohort who received doula care through MOMCares between January of 2022 and June of 2024 compared to the population at large and the overall cohort, and rates were lowest among those who attended 100% of their prenatal appointments. The c-section rate among the cohort members who received doula care was 20%, and as low as 18.52% among those who attended all their prenatal appointments; this is slightly lower than the rate of 24.44% among the cohort at large and lower than the rate of 36.51% among the PRAMS Maryland group. Preterm delivery rates among the MOMCares doula patients were as low as 7.69% for those who attended all appointments; again, significantly lower than the preterm delivery rate of 15.09% among the MOMCares cohort and 22.73% among the Black members of the cohort. As for low-birth-weight deliveries, they occurred at a rate of 3.33% among doula patients who attended all appointments, compared to 7.50% among the MOMCares cohort and 14.29% among the Black members of the cohort.

These sizable differences suggest that not only does prenatal and continuous labor support through a doula reduce the risk of adverse birth outcomes, but the risk also decreases further as clients attend more of their prenatal appointments and take part more fully in the program. Although the size of this sample yielded *p* values (in Fishers Exact Tests) below 0.05, which did not allow us to reject the respective null hypotheses (that the variables in question are not associated or related), extant research performed with larger, and more diverse samples, suggest that these variables are indeed associated.

March of Dimes reports as recent as 2023 show that c-section delivery rates continue to be higher among Black birthing people, a disparity we saw reflected in the PRAMS phase 8 data for Maryland. A 2015 study highlights this disparity and shows that even after risk adjusting for various other factors contributing to c-section delivery, Black birthing people still are 1.27 times more likely to experience a c-section delivery than their white counterparts [13]. Most recent March of Dimes data also shows that Black infants are approximately 2 times more likely to be born low birthweight than white infants [3], a disparity also highlighted in a study examining infants of various racial backgrounds born between 1999 and 2014. This study showed that while Black infants were more likely to be born with low birth weight, they were less likely to be born with macrosomia – defined as a birthweight above 4000g [14].

Several studies have highlighted the size of the disparity in preterm birth rates between Black birthing people and other racial groups, showing that Black birthing people have the highest rate of preterm birth in the United States, while white birthing people have the lowest rate. The most recent of these studies, published in 2024, examined 34 million live births in the US, and suggested that non-US-born birthing people may have lower rates of preterm delivery – no matter their race – when compared to their US counterparts [15]. In studies that examine delivery methods in relation to preexisting maternal medical conditions or morbidities such as diabetes, hypertension, and previous c-section deliveries, it has been found that c-sections occur in as many as 50% of deliveries to birthing people with severe maternal morbidity [16].

Within the context of this existing, and more extensive, research, the data from this analysis suggests that birthing people who received doula care from MOMCares benefited, in the form of fewer negative or poor birth outcomes, from the support. Furthermore, one qualitative question was asked in our survey – “If you received doula care services, do you feel that it contributed to a more culturally competent and positive birth experience?” 80% of those who responded to this question indicated “yes.” For clarity, only one person responded “no,” while one person responded that this question was not applicable to them (likely a birthing person who did not receive doula care services through MOMCares), and one person responded that they applied for services late in pregnancy and were assigned a doula on the day of delivery, and so this care did not contribute to their overall pregnancy experience.

## V. Conclusion and Implications

This study and its results contribute to a rapidly growing body of research which supports the theory that continuous labor support and advocacy, provided through a trained doula, improves birth outcomes. This improvement is particularly significant in Black and other minority populations, as well as among those with preexisting medical conditions or high-risk pregnancies. Furthermore, doula care may contribute to a more positive pregnancy and birth experience as well as more culturally competent care. This speaks to the need for better diversity and equity training for OBGYN providers, as well as patient advocates, but supports the need for doula care as an accessible and affordable service for minority birthing people.

## VI. Limitations

Despite the strengths of this study, several limitations should be acknowledged. First, the sample size of 124 birthing people, while adequate for descriptive analysis, limits the generalizability of the results to a broader population.

Furthermore, data for each variable was not available for each member of the cohort, which made subsamples smaller for some comparisons and analyses. The study participants were primarily from the Baltimore region and received intake for doula care through MOMCares, MD, but did not all receive doula care, and may not be representative of all birthing people receiving doula care in different geographic locations or through other programs. PRAMS data from the Maryland site was used for comparison, but the cohort for this research is significantly less racially diverse than the PRAMS sample. Second, reliance on self-reported data for variables such as maternal health concerns and socioeconomic status may introduce response bias. The low response rate of 12% to the survey may introduce a non-response bias as well, though MOMCares provided data for most intake clients initially.

Additionally, potential confounding variables like detailed medical histories and environmental factors were not accounted for in this study. Some medical factors could influence both maternal health and birth outcomes but have been considered in earlier research and should continue to be considered in future research.

## Data Availability

All data produced or analyzed during this study are included in this published article. The primary community partner dataset (MOMCares) contains sensitive participant information and is available from the corresponding author upon reasonable request and with permission of MOMCares, MD. Secondary Pregnancy Risk Assessment Monitoring System (PRAMS) data are available through the Centers for Disease Control and Prevention (CDC) and the Maryland Department of Health upon application and approval of a formal Data Use Agreement.

## VII. Acknowledgement

We thank the PRAMS Working Group, which includes the PRAMS Team, Division of Reproductive Health, CDC, and the Maryland PRAMS site for their role in conducting PRAMS surveillance and allowing the use of their data.

## Notes

### Competing Interest Statement

The authors have declared no competing interest.

### Author Declarations

Ethics committee of the Association of Maternal and Child Health Programs, Graduate Student Epidemiology Program gave ethical approval for this work.

